# 16S rRNA gene and *rpoB* Nanopore Sequencing for Bacterial Detection and Identification in Clinical Samples

**DOI:** 10.1101/2025.09.30.25336965

**Authors:** Joanna Małgorzata Bivand, Audun Sivertsen, Marit Gjerde Tellevik, Oystein Saebo, Robin Patel, Ruben Dyrhovden, Øyvind Kommedal

## Abstract

Broad-range amplification and sequencing of the 16S ribosomal RNA (rRNA) gene directly from clinical samples can improve diagnosis of bacterial infections. Recent advances in the Oxford Nanopore Technologies (ONT) third-generation sequencing platform, delivering sequencing accuracy exceeding 99%, makes it a possible option for this approach. However, even full-length 16S rRNA sequencing may lack resolution to distinguish closely related species, a problem exacerbated by the remaining ONT error rate.

Here, we propose a strategy combining ONT sequencing of the 16S rRNA gene with a segment of the *rpoB* gene. We further developed a novel bioinformatics tool mitigating issues related to the ONT error rate, and also providing homology scores allowing for implementation of CLSI interpretive criteria. Using this approach, we investigated 25 abdominal abscess samples, with Illumina sequencing of the same genes as the comparator method. Our results showed that *rpoB* ONT sequencing provided species-level identification in 91.5% of detections, as compared to 68.9% with 16S rRNA ONT sequencing. However, 16S rRNA ONT sequencing showed the highest sensitivity detecting 84.0% of the total identifications as compared to 74.2% for *rpoB*. Together, 16S rRNA and *rpoB* ONT sequencing achieved a detection rate of 94.0%, with species-level identification in 87.7%. For both gene targets, ONT sequencing paralleled Illumina sequencing. Combining ONT sequencing with optimized bioinformatics processing, alongside the high sensitivity of 16S rRNA broad-range amplification and the improved taxonomic resolution of *rpoB*, enable accurate detection and identification of bacteria directly from clinical samples.

## INTRODUCTION

Broad-range amplification of the bacterial 16S ribosomal RNA (rRNA) gene directly from clinical samples followed by next-generation sequencing has highlighted shortcomings of bacterial culture, particularly for samples containing anaerobic or fastidious microbes and for those from patients who have received antibiotics prior to sample collection (1–5). Despite its potential, the adoption of 16S rRNA targeted next-generation sequencing (16S TNGS) in clinical microbiology has been slow, mainly due to high cost and long turn-around time. Although the price per sequenced base has dropped over time, next-generation sequencing platforms are not optimized for the needs of diagnostic microbiology, which demand small and frequent runs with short turn-around times. Oxford Nanopore Technologies (ONT) sequencing is a rapid, long-read sequencing technology offering smaller, single-use flow cells (Flongle flow cells) with a cost and capacity potentially suitable for analysis of a single sample. A higher sequencing error rate and lack of appropriate bioinformatics tools have prevented widespread adoption of this platform in diagnostic microbiology. Today, with a read accuracy exceeding 99% combined with bioinformatics approaches prioritizing accuracy over real-time analysis of data, ONT sequencing is beginning to yield results of sufficient quality for infectious diseases diagnostics (6–8).

A limitation of 16S TNGS is poor species-level resolution within some clinically important genera (9). This problem is exacerbated by the increasingly fine-grained bacterial taxonomy where novel species are often the result of splitting of previous polyphyletic taxa into two or more closely related species (10–12). Even full-length 16S rRNA gene sequencing will not uniformly resolve these challenges and the actual improvement in classification as compared to the partial 16S rRNA V1-V3 region is debatable (6, 8, 13, 14). *rpoB* is an alternative marker gene with significantly better resolution (15–17). Recently, we described the first broad-range *rpoB* primers suitable for bacterial detection and identification in clinical microbiology (18). The resulting ∼550 base pair (bp) amplicon is well-suited for ONT sequencing.

According to CLSI guidelines for bacterial identification based on the 16S rRNA gene, unambiguous species-level identification generally requires ≥99% homology with a high-quality reference and a distance to the next alternative species of >0.8% (19). Similar criteria have been published for *rpoB*, recommending a homology to a high-quality reference of ≥98.5% and a distance to the next alternative species >1.4% (18). Such criteria reflect the phylogenetic properties and biological variation of the respective genes and should be used regardless of sequencing method. However, their application is dependent on an alignment and cannot be coupled with pure k-mer based identifications. To address this, we collaborated with a bioinformatics company (Pathogenomix; Santa Cruz, CA) to develop a novel module in the RipSeq software optimized for analysis of ONT amplicon sequencing data based on binning of similar reads followed by a consensus sequence generation from each bin and ultimately, identification using BLAST.

The aim of the present study was to evaluate targeted Nanopore sequencing of partial 16S rRNA and *rpoB* genes for bacterial detection and identification directly from clinical samples using Illumina sequencing of the same target genes as the comparator method. The investigation included (i) Nanopore sequencing of partial *rpoB* (550 bp) and 16S rRNA amplicons from 25 intraabdominal samples and a commercial staggered mock community; (ii) design of a novel broad-range *rpoB* forward primer to pair with our previously published broad-range *rpoB* reverse primer, generating a shorter ∼450 bp amplicon suitable for Illumina sequencing and Illumina sequencing of partial *rpoB* and 16S rRNA amplicons from the same samples; (iii) development of a novel software for analysis of ONT amplicon sequencing data; (iv) creation of a *rpoB* gene database; (v) comparison of results obtained by ONT and the novel bioinformatics tool with those from the established Illumina pipeline; and (vi) assessment of the diagnostic value of *rpoB* compared to 16S rRNA gene sequencing for the Illumina and Nanopore platforms.

## MATERIALS AND METHODS

### Ethical statement

The study was approved by the Regional Committee for Medical and Health Research Ethics for the western region, Norway (REK 2024/728299).

### Biological samples and controls

#### Clinical samples

Remnant extracted DNA from 25 abdominal abscess samples, previously investigated with 16S rRNA gene PCR and Sanger sequencing in routine clinical practice, were studied. Extraction of bacterial DNA was performed as described previously (20); the extracted DNA had been stored at -80°C at the Department of Microbiology, Haukeland University Hospital, Bergen, Norway.

#### Mock community samples

The commercial mock community (ZymoBIOMICS® Gut Microbiome Standard, Microbiomics, Zymo Research, Irvine, California, USA) was diluted 1:10 and 1:100 in nuclease free water. The dilutions were further diluted 1:2 in MagNa Pure Bacterial Lysis Buffer (BLB) (Roche, Basel, Switzerland) and homogenized using a FastPrep®-24 instrument (MP Biomedicals, Irvine, California, USA). Extraction of bacterial DNA was performed using the ELITe InGenius automated extractor with the ELITe InGenius SP 200 Extraction Kit (ELITechGroup MDx, Turin, Italy) following the manufacturer’s protocol.

#### Controls - Nanopore sequencing

Negative controls consisting of BLB and PCR-grade water were extracted together with clinical samples and included with each PCR setup. Seven extraction controls for 16S rRNA gene and four extraction controls for *rpoB* were sequenced separately on separate Flongle flow cells.

#### Controls - Illumina sequencing

Positive (*Yersinia enterocolitica* ATCC 23715) and negative (BLB and PCR grade water) extraction controls were included with each sequencing run.

### Amplification of partial 16S rRNA and *rpoB* genes

#### PCR Primers

Primers are listed in Table 1. The *rpoB* primers and 16S V1-V3 primers used for Nanopore sequencing have been described and evaluated previously (18, 20, 21). The 16S rRNA primers used for Illumina sequencing are those recommended in the standard Illumina protocol (22), with modifications as described previously (21). For design and evaluation of the novel Illumina *rpoB* forward primer, reference sequences from the NCBI database were aligned in Geneious bioinformatics software version 9.1.7 (Dotmatics, Auckland, New Zealand). Combined with our previously described *rpoB* reverse primer (18) the resulting Illumina *rpoB* amplicon corresponded to *rpoB* nucleotide positions 1,675 to 2,068 of the *Escherichia coli* ATCC 11775 type strain, with amplicon sizes ranging from 388 bp to 445 bp depending on the species. Universality of the primer was confirmed *in silico* against bacteria representing 124 clinically relevant genera.

**TABLE 1.**
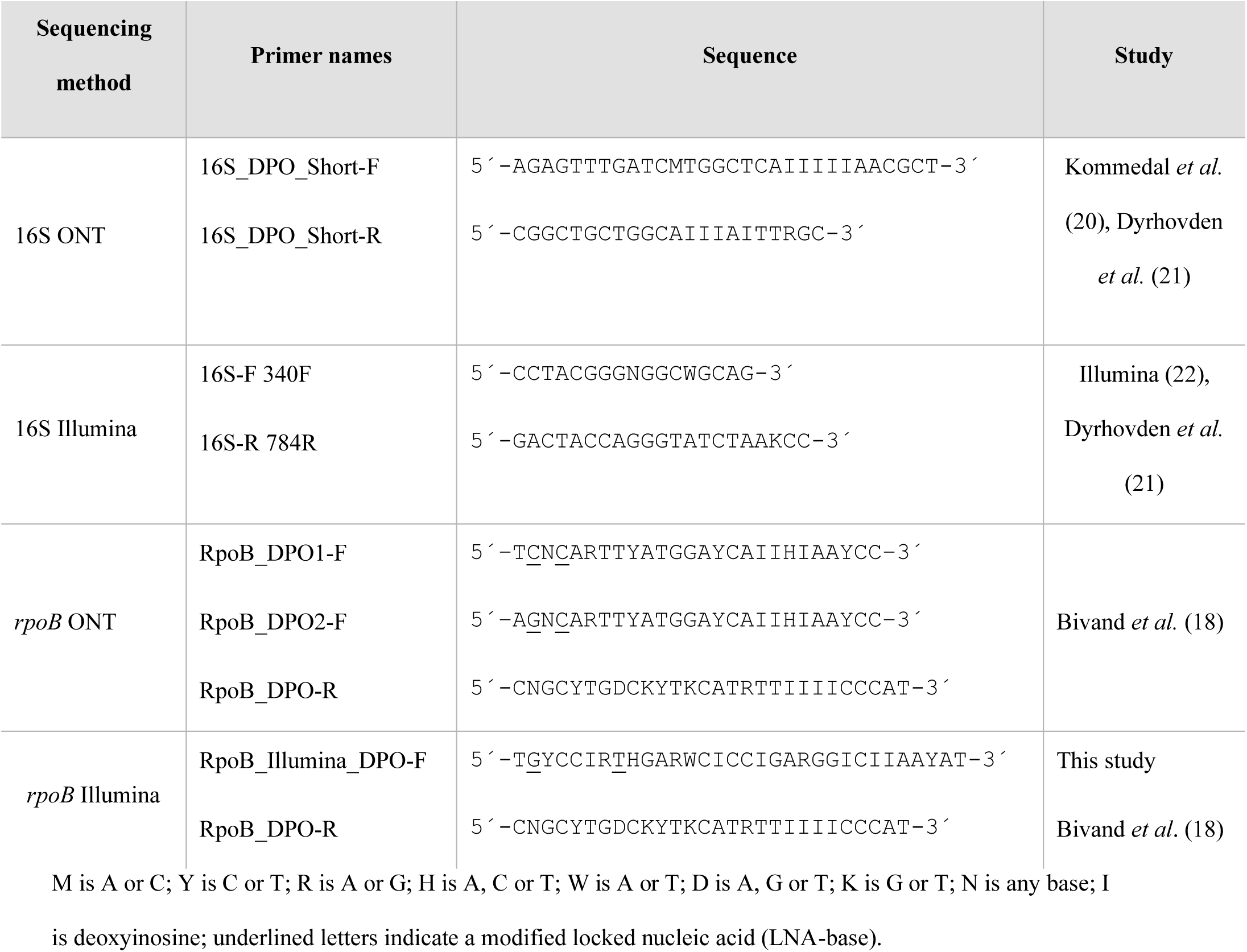
16S rRNA and *rpoB* primers for ONT and Illumina sequencing.

In addition, as a supplementary evaluation of the coverage of *rpoB* primers, an extensive *in silico* PCR analysis was conducted using the Usearch11 software (https://www.drive5.com/usearch/manual/cmds_search.html) and the Genome Taxonomy Database (GTDB). Mismatches in primer 3’-end believed to impair amplification and other significant mismatches for clinically relevant species are displayed in Supplementary Table S1.

#### PCR conditions

The real-time PCR assays were carried out using a QuantStudio™5 Real-Time PCR System (Thermo Fisher Scientific). The 16S rRNA and *rpoB* PCRs for Nanopore sequencing were run as previously described (18). Amplification of the 16S rRNA V3-V4 region was performed as previously described (21). For Illumina, the *rpoB* PCR mixture (25 µl) contained 12.5 µl TB Green Premix ExTaq (Takara Bio, Kusatsu, Japan), 4 µl forward primer, 3 µl reverse primer (both from 10 µM solutions), 3.5 µl PCR grade water, and 2 µl of template DNA. The PCR thermal profile included an initial denaturation at 95°C for 30 s, 45 cycles of denaturation at 95°C for 20 s, annealing at 50°C for 20 s and elongation at 72°C for 30 s.

### Sequencing

#### ONT sequencing

Library preparation was performed using the Ligation Sequencing Kit V14 (SQK-LSK114) and the protocol Ligation Sequencing Amplicons V14 protocol (ONT). Samples were sequenced on the GridION platform using R10.4.1 Flongle flow cells with a sequencing duration of 6 hours for all samples. Sequencing was performed using super-accurate basecalling and minimum Q-score of 10. Only Flongle flow cells with more than 50 active pores were used.

#### Illumina sequencing

The library was prepared according to the Illumina 16S Metagenomic Sequencing Library Preparation protocol with minor modifications (21); sequencing was performed using the MiSeq Reagent Kit v3 (2 x 300 bp) on the Illumina MiSeq System (Illumina, Redwood City, CA).

### Interpretative criteria and data analysis

#### Taxonomy criteria and definition of a valid identification

For taxonomic assignments, three classification levels were used: 1) Species-level: ≥99% (16S)/≥98.5% (*rpoB*) identity with a reference sequence, combined with a ≥0.8% (16S)/≥1.4% (*rpoB*) distance to the next closest species. 2) Species-group: identification meeting the species-level homology threshold, but with distance to the next closest species below 0.8% (16S) or below 1.4% (*rpoB*). 3) Genus-level: when the homology score was below the species-level criteria but ≥97%. For contaminant filtering, the sample-specific cutoff criteria described by Dyrhovden *et al.* (23) were applied. The filtering principle was developed and evaluated specifically for clinical microbiology. All non-contaminant detections were considered valid, including those made by only one method.

#### Reference databases

The primary 16S rRNA reference database was the “Pathogenomix Prime 16S” (Pathogenomix) database containing 3000 manually curated complete gene references, supplemented with (i)16S rRNA references extracted from all whole genome references in GenBank; (ii) GenBank 16S rRNA references linked to a scientific publication; (iii) all 16S rRNA references included in the “The All Species Living Tree” Project (https://imedea.uib-csic.es/mmg/ltp/); and (iv) all 16S rRNA references from the Human Oral Microbiome database (https://homd.org/). The *rpoB* database was derived from the GTDB database (https://gtdb.ecogenomic.org/). GTDB has implemented a standardized microbial taxonomy based on genome phylogeny. The GTDB taxonomy uses reference strains as fixed points for nomenclature and classification, and ANI distance (>95%) of circumscribed species to representative genomes (24). This results in clusters forming around type strain genomes with >95% ANI. Unsuffixed names represent clusters or lineages that contain a type strain, whereas an alphabetic suffix at the end of a species name indicates a placeholder name for a cluster without a type strain, suggestive of a novel, undescribed species. To construct a *rpoB* database working with RipSeq, bac120 (v.r226) representative sequences were downloaded from GTDB (25), and a multiple FASTA file containing *rpoB* sequences (TIGR02013.fna) was extracted.

#### Sequence analysis - Illumina

FASTQ-files were individually analyzed using the NGS module in the RipSeq software (Pathogenomix) (1, 4, 21, 23, 26). Briefly, sequencing reads shorter than 300 bp and any remnant primer sequences were removed before *de novo* clustering of reads into operational taxonomic units (OTUs) using homology thresholds of 99.0% for the 16S rRNA gene and 98.5% for *rpoB*. A representative sequence from each OTU was subsequently identified using a standard BLAST-search against the “Pathogenomix Prime 16S” database or the novel GTDB-based *rpoB* database (supplemented with searches against relevant NCBI GenBank database if only a low homology score was obtained with the standard databases).

#### Sequence analysis - ONT

FASTQ-files were analyzed individually using the novel Long Read Sequencing (LRS) module optimized for ONT in the RipSeq software. Reads shorter than 450 bp for 16S rRNA gene or 500 bp for *rpoB*, and any remnant primer sequences were trimmed before k-mer based read binning against the reference-databases for 16S rRNA gene (GenBank RefSeq 16S database) or *rpoB* (the novel GTDB-based database). Subsequently, consensus sequences were generated based on multiple alignments of reads that had binned to the same reference (consensus sequence generation). Finally, consensus 16S rRNA gene and *rpoB* sequences from each bin were identified using a BLAST search against the “Pathogenomix Prime 16S” or the novel GTDB-based *rpoB* database, respectively (supplemented with searches against relevant NCBI GenBank database if only a low homology score was obtained with the standard databases).

### Statistical analysis

All non-contaminant detections were considered valid, including those made by only one method. Statistical analysis was performed in R v 4.3.2. The phyloseq package (v1.46.0) was used to calculate alpha-diversity metrics, including Chao1 and Shannon indexes, and to estimate richness by method. *P* values were adjusted using the Kruskal-Wallis test, with a threshold of *P* value of <0.05 indicating statistically significant differences. Concordance between methods was assessed based on the number of observed OTUs. The ggplot2 package (v3.5.2) was used for data visualization.

### Data availability

The 16S rRNA gene and *rpoB* sequences generated in this study will be deposited in the European Nucleotide Archive (ENA) at European Molecular Biology Laboratory-European Bioinformatics Institute (EMBL-EBI). We are in the process of submitting the data.

## RESULTS

The 16S rRNA gene and *rpoB* amplicons from 25 intraabdominal abscess samples and one commercial staggered mock community were successfully sequenced using ONT and Illumina sequencing platforms (Figure 1). The average number of reads per sample was 368,626 for ONT and 610,080 for Illumina (Supplementary Table S2, Supplementary Figure S1). Twenty samples, including the mock community, were polymicrobial, with diversities spanning from two to 38 bacteria. In total, there were 318 bacterial detections, representing 91 genera and 172 species. Among these, 55 (17.3%) were made by only one method, 47 (14.8%) by two methods, 58 (18.2%) by three methods and 158 (49.7%) by all four methods. Detailed sequencing results for each sample are provided in Supplementary Table S3.

**FIG 1.**
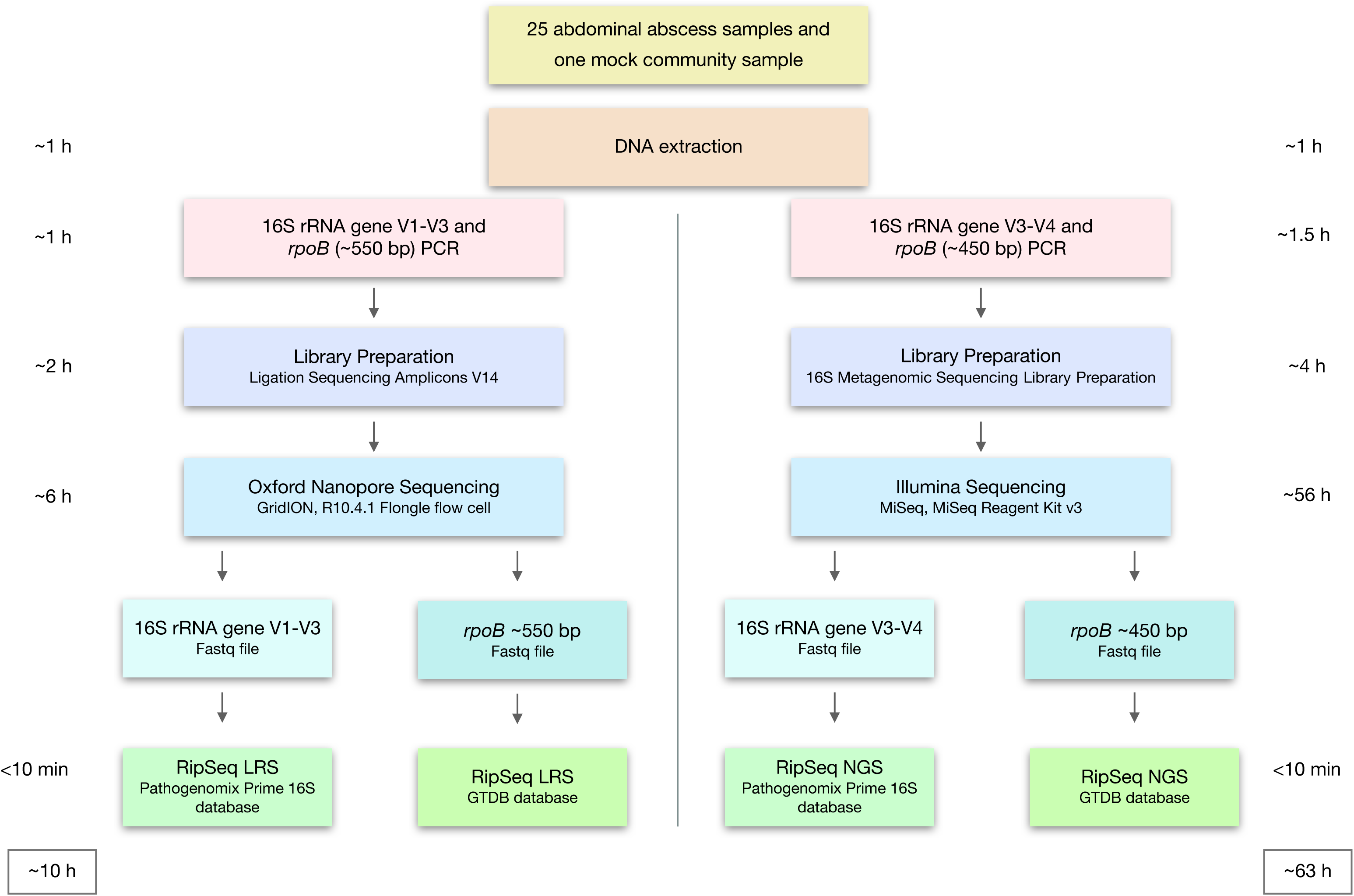
Overview of experimental workflow. Turn-around times for analyses are shown on the sides. Time to results is indicated in the boxes at the bottom of the figure.

### Sensitivities and taxonomy classification level

The *rpoB* amplicons provided the best taxonomic resolution. *rpoB* ONT enabled species-level classification for 91.5% and *rpoB* Illumina for 86.5% of bacteria detected by each method (Figure 2c). In comparison, 16S rRNA gene ONT and 16S rRNA gene Illumina allowed for identification to the species-level for 68.9% and 56.1% of detections, respectively. 16S rRNA gene ONT sequencing was the most sensitive method and detected 84.0% of the total microbial detections whereas *rpoB* Illumina was the least sensitive method and detected 67.6% (Figure 2a).

**FIG 2.**
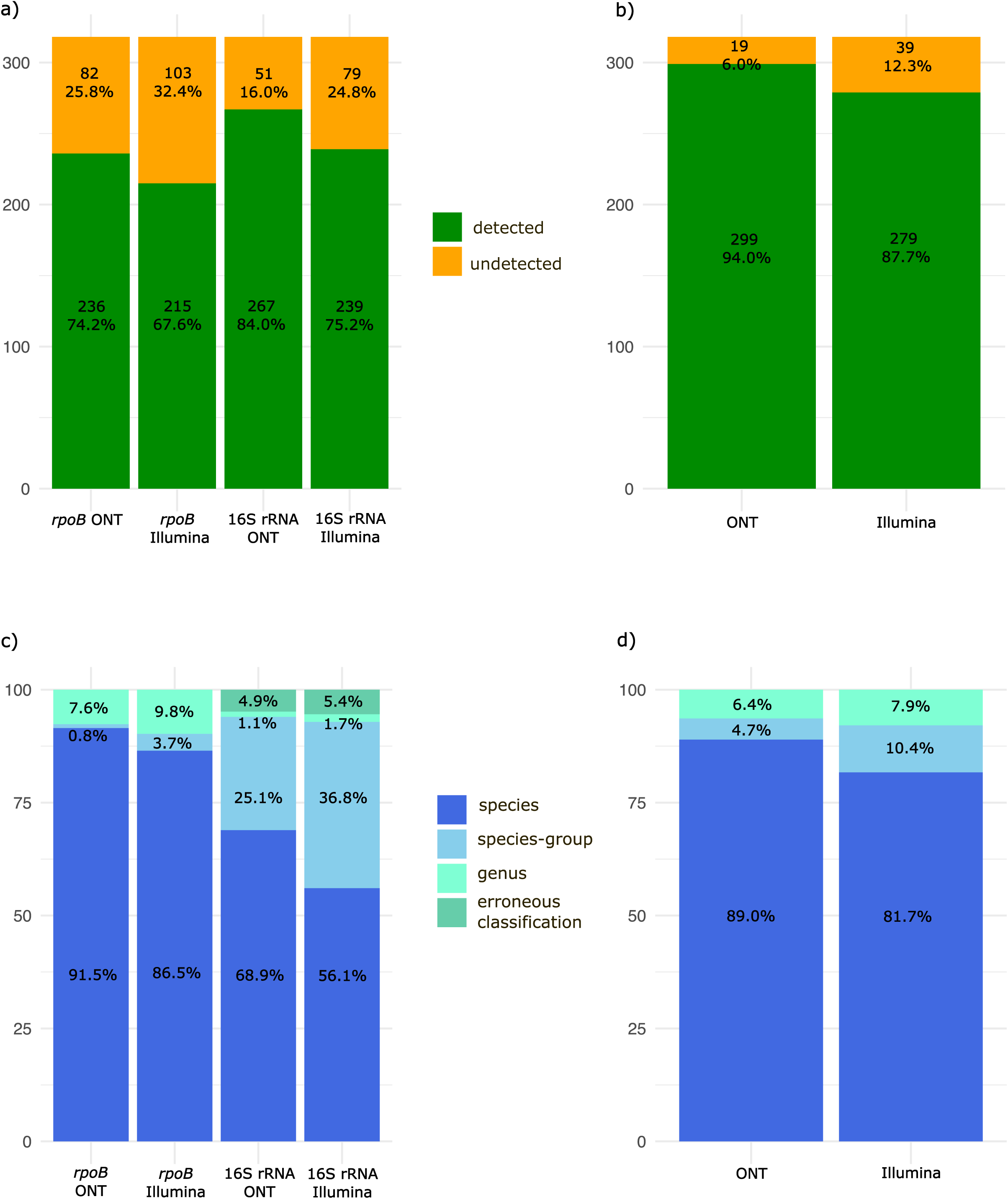
Results from 16S rRNA gene and *rpoB* ONT and Illumina sequencing. (a) Distribution of detected and undetected bacteria for *rpoB* ONT, *rpoB* Illumina, 16S rRNA gene ONT and 16S rRNA gene Illumina. (b) Theoretical estimation of detected and undetected bacteria using combination approaches: (i) *rpoB* ONT + 16S rRNA gene ONT and (ii) *rpoB* Illumina + 16S rRNA gene Illumina. (c) Taxonomic assessment of detected bacteria at three classification levels: species, species-group, and genus. Bacteria likely misclassified to species or species-group levels by16S rRNA gene sequencing are highlighted in light green. d) Theoretical estimation for distribution of taxonomic classification levels for combined approaches.

Combining *rpoB* and 16S rRNA gene for describing polymicrobial infections may represent an attractive approach for maximizing sensitivity and taxonomic resolution. The estimated detection rates for *rpoB* and 16S rRNA gene combined were 94.0% for ONT and 87.7% for Illumina, with species-level identification achieved for 89.0% and 81.7%, respectively (Figure 2b, d).

Taxonomic resolutions for detections made by all four sequencing approaches are compared in Figure 3a. In the present study, *rpoB* improved resolution within the genera *Acinetobacter*, *Aeromonas*, *Campylobacte*r, *Edwardsiella*, *Eikenella*, *Enterobacter*, *Enterococcus*, *Fusobacterium*, *Klebsiella*, *Staphylococcus*, *Stenotrophomonas*, *Streptococcus* and *Veillonella*. The shorter *rpoB* amplicon used for Illumina sequencing performed similarly to the *rpoB* ONT amplicon, with a slightly lower resolution for *Bacteroides* and *Enterobacter*. The 16S rRNA gene V3-V4 amplicon used for Illumina sequencing provided the lowest resolution and was inferior to the 16S rRNA V1-V3 region for multiple genera, most notably *Fusobacterium*, *Streptococcus* and *Bacteroides*. The poorer resolution of the 16S rRNA gene resulted in the misclassification of several possibly novel species (Supplementary Table S4). The most frequent example was five valid identifications of *Campylobacter gracilis* by 16S rRNA gene analysis, where the corresponding *rpoB* sequences obtained only 92.8%-94.6% homologies with the *C. gracilis* type strain, pointing to a novel *Campylobacter* species. The 16S rRNA gene ONT was the only method differentiating between *Bacteroides uniformis* and *Bacteroides rodentium*, representing the sole instance where the 16S rRNA gene provided better discrimination than *rpoB*.

**FIG 3.**
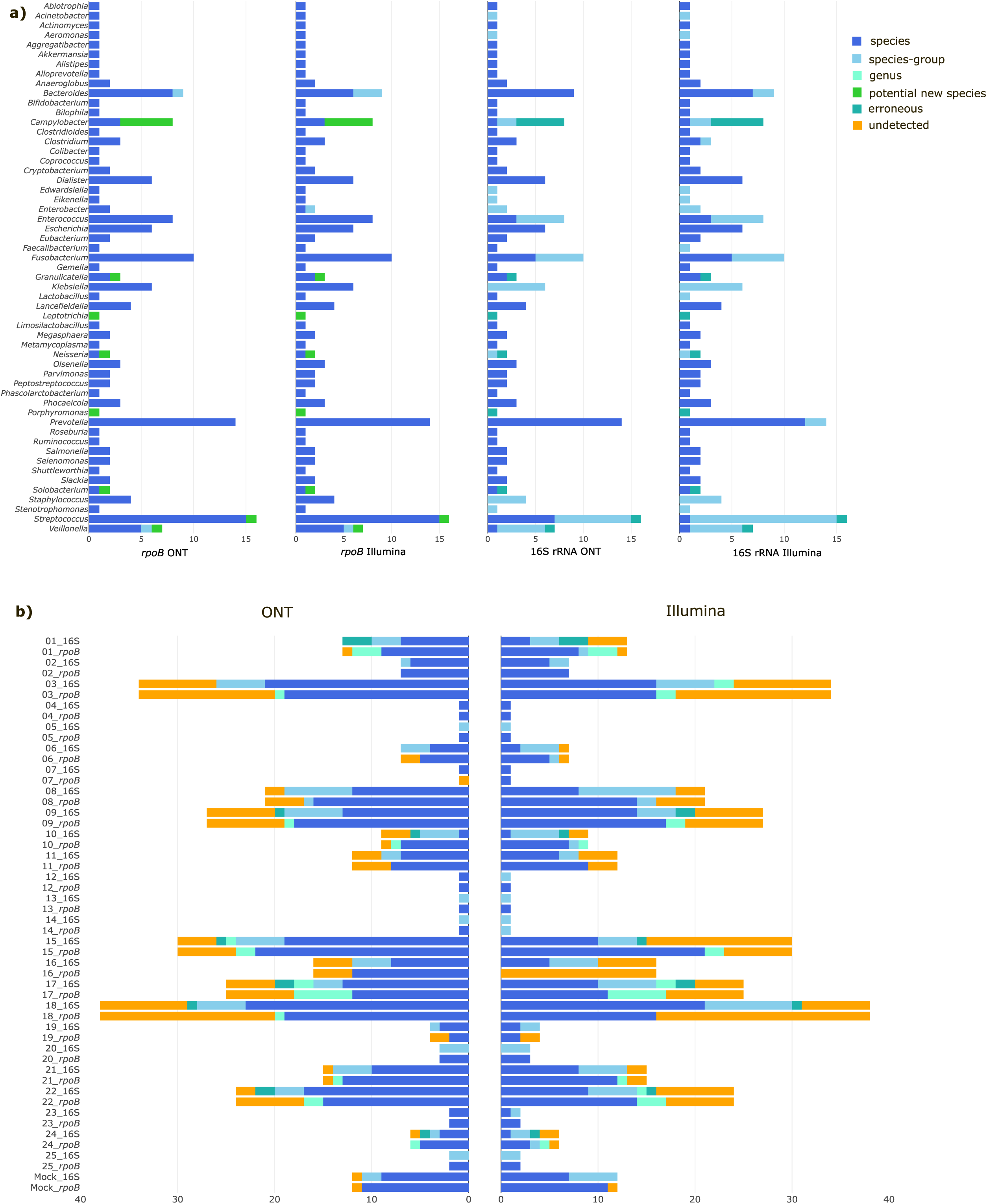
Taxonomic classification. (a) Taxonomic classification for detections made by all four methods across all samples. Numbers below the figure indicate the number of detected bacteria within each genus. Bacteria likely misclassified to species or species-group by 16S rRNA gene sequencing are highlighted in dark green (erronouse). Bacteria representing potential novel species are highlighted in light green. (b) Overview of ONT and Illumina sequencing results for the 16S rRNA and *rpoB* genes across all samples. Sample IDs are shown on the y-axis. Numbers below the figure indicate total microbial detections.

### Sample characterizations

Among the polymicrobial samples, complete descriptions by all four methods were achieved for only four (S2, S20, S23 and S25). In addition, complete descriptions were achieved by 16S rRNA gene ONT for three samples (S1, S6 and S19), for two samples by 16S rRNA gene Illumina (S19 and the mock community) and for one each by *rpoB* ONT (S24) and *rpoB* Illumina (S10) (Figure 3b). In general, the number of undetected species correlated with increased sample complexity. In ten samples, no methods detected all pathogens; these were complex samples with at least twelve, and in most cases over twenty species detected (range 12 to 38).

The six monomicrobial samples were correctly described by all methods except for sample S7 where *rpoB* ONT failed to detect *Actinomyces israelii*, also represented by a low number of reads with *rpoB* Illumina. This is most likely explained by the extremely high GC content (71.2%) of the *A. israelii rpoB* amplicon resulting in poor target amplification and more nonspecific amplification of other DNA in the sample.

### Background contaminants

The most abundant and frequently detected contaminants used to set the sample specific cutoffs were *Cutibacterium acnes*, *Sphingomonas echinoides* and *Staphylococcus epidermidis* for *rpoB* ONT and *C. acnes* for 16S rRNA gene ONT. Presumed contaminants filtered from the clinical samples and species detected in the negative controls are listed in Supplementary Table S5.

### Missed bacterial detections

The highest proportion of undetected bacteria was for *rpoB* Illumina (32.4%) and the lowest for 16S rRNA gene ONT (16%) (Figure 2 a). Some missed detections were explained by signifi cant primer mismatches: The *rpoB* ONT forward primer has a mismatch with *Corynebacterium*; the *rpoB* ONT reverse primer has a mismatch with *Lactobacillus*; and the 16S rRNA gene ONT forward primer has a mismatch with *Bifidobacterium*. Both *rpoB* primer pairs and the 16S rRNA gene ONT primers have multiple mismatches with the Archaea *Methanobrevibacter smithii* included in the mock community. *Schaalia odontolytica* was not detected by *rpoB* sequencing despite perfect primer matching, even though it was reported at relatively high concentrations by 16S rRNA gene sequencing (S17). Presumably, the high GC content of the *rpoB* gene (∼65%) of *Schaalia* species resulted in this amplicon being outcompeted in the PCR by *rpoB* amplicons with lower GC contents from other species in the same samples. That said, the majority of missed detections occurred among low level identifications in complex polymicrobial samples. In broad-range amplification and sequencing, low-abundance bacteria are prone to being outcompeted during PCR or sequencing, and their detection may depend upon variations in primer-binding or amplification efficacy, differences in sequencing depth or chance (23).

### Potentially novel species

Twelve bacteria with a GTDB placeholder name (suffix at the end) and *rpoB* homology scores >98.5% were identified. Due to the absence of an established taxonomy for these species, the current GTDB classification was used, and they were reported at the species-level with a notification that the taxonomy may change. Using the criterion that a *rpoB* homology <97% against any reference available in GTDB/GenBank is indicative of a new species, 18 candidate novel species were identified (Supplementary Table S4). Twelve of these are also indicated in Figure 3a.

### Concordance between methods

To compare microbial diversities identified by the four sequencing approaches, Chao1 and Shannon alfa diversity analysis were performed (Supplementary Figure S2 and Supplementary Figure S3). The highest richness was obtained by 16S rRNA gene ONT followed by *rpoB* ONT (Figure 4e). However, using Kruskal-Wallis test, there were no significant differences between methods (P = 0.86). Concordance between methods, as presented in Figure 4a-d, shows good correlation between 16S rRNA gene ONT and *rpoB* ONT sequencing results, and between *rpoB* ONT and *rpoB* Illumina sequencing results. In contrast, the correlation is lower for 16S rRNA gene Illumina *versus rpoB* Illumina, and for 16S rRNA gene Illumina *versus* 16S ONT.

**FIG 4.**
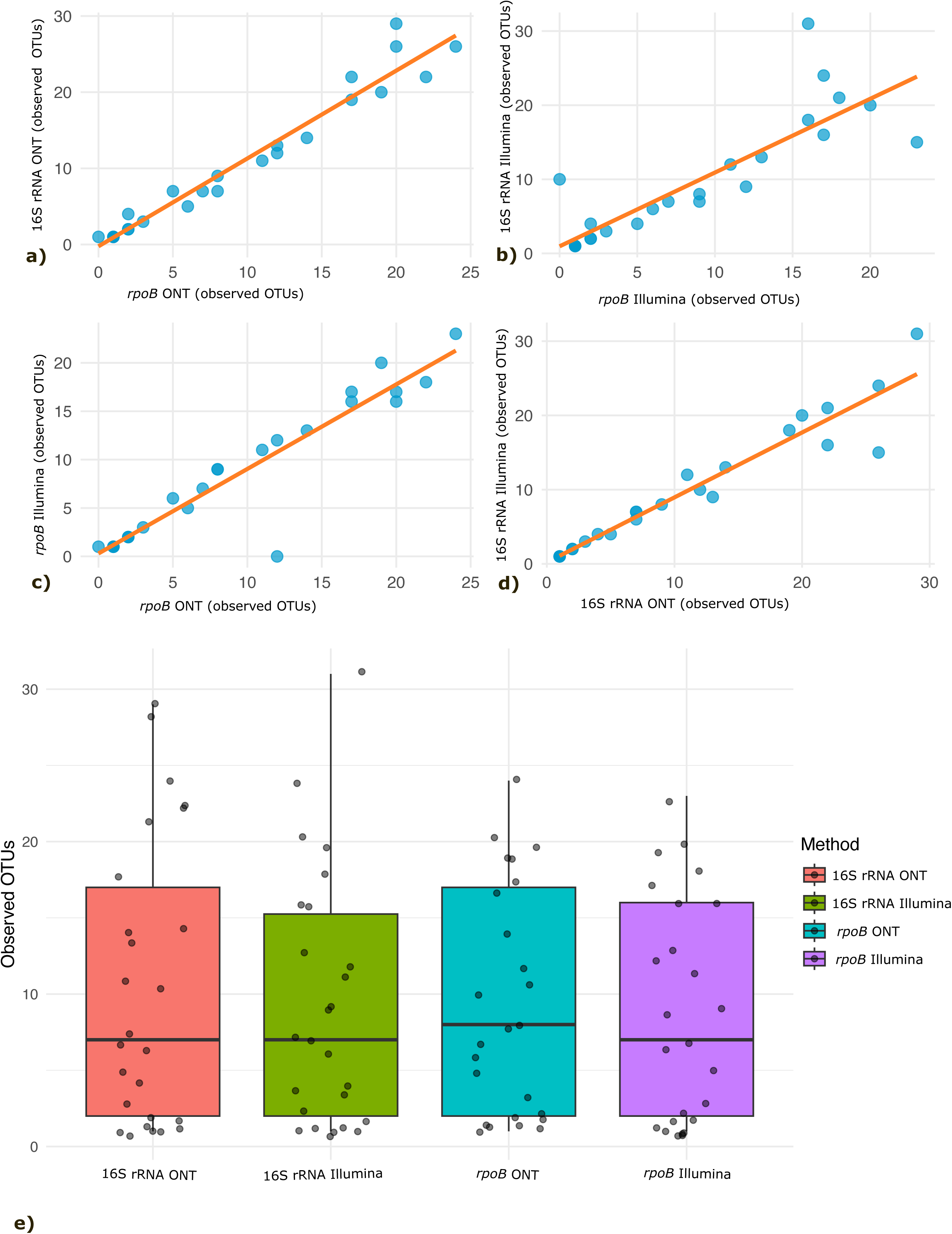
Concordance between methods based on the number of observed OTUs. (a) Concordance between 16S rRNA gene ONT and *rpoB* ONT. (b) Concordance between 16S rRNA gene Illumina and *rpoB* Illumina. (c) Concordance between *rpoB* Illumina and *rpoB* ONT. (d) Concordance between 16S rRNA gene Illumina and 16S ONT. (e) Observed richness measured across all four sequencing approaches.

## DISCUSSION

Here, we present and evaluate a complete approach for Nanopore sequencing of bacterial phylogenetic marker genes directly from patient samples in clinical microbiology. We used a novel bioinformatic tool to optimize data interpretation and included broad-range amplification and sequencing of the *rpoB* to mitigate limitations in the taxonomic resolution of the 16S rRNA gene.

The total preparation time from DNA extraction to the start of ONT sequencing was 4-5 hours, comparable to the preparation time for Sanger-sequencing currently in use in many laboratories. Although this will increase somewhat with the number of samples, sequencing can still be started the same working day, providing next day results. Bioinformatics processing time was 3-8 minutes per sample depending on sequencing depth and sample complexity. Each sample was individually sequenced, and separate Flongle flow cells were used for 16S rRNA gene and *rpoB* sequencing. However, the Flongle output should be sufficient to sequence both gene targets on the same cell, separating them either by barcoding or bioinformatically during the identification process. With overnight sequencing, the output might even allow for multiplexing of 2-4 samples. For laboratories with larger sample volumes, or if some batching of samples is acceptable (e.g., sequencing twice a week), a more cost- and time-efficient approach would be to sequence all samples and both targets simultaneously on a larger MinION flow cell using barcoding.

For filtering of background contamination, the approach described by Dyrhovden *et al.,* in which the dominant contaminant species are used to establish sample-specific cutoffs, was applied (23). As the dominant background bacterial DNA contamination appeared stable, it might be sufficient to run the negative controls once or twice per week and with new lots of reagents or disposables. Multiplexing samples on a MinION flow cell will allow for inclusion of controls with each run.

For bacterial identification based on broad-range amplification and Sanger sequencing of the 16S rRNA gene, the CLSI recommends using the 500 bp V1-V3 segment, considered the most variable part of the gene (19). For species level discrimination, full-length 16S rRNA sequencing enabled by ONT has been framed as an improvement (6, 8, 13, 14). However, for alignment-based approaches applying CLSI interpretive criteria, it is not intuitive that full-length 16S rRNA sequencing will improve species-level discrimination, since including less variable regions may reduce the percentage difference to the next alternative species (27). For Bayesian classifiers, to theoretically exploit any statistically significant difference between neighboring species, this might be different. A study using such a classifier suggests that full-length 16S rRNA gene sequencing provides species level resolution and outperforms partial 16S rRNA analysis (28). However, this *in silico* analysis was limited to a preselected subset of 16S rRNA gene sequences that were all ≥1% different from each other over the full-length gene, assuring 100% species-level resolution. The study design further biased comparisons with alternative partial segments, since references differing by more than 1% in a subsegment were excluded if they showed less than 1% difference across the entire gene. Even in this comparison, the V1-V3 segment achieved 90% species-level resolution, and the authors suggested that highly informative 16S rRNA gene regions, such as V1-V3, may be sufficient for species-level discrimination. In a later study, it was reported that between 14 and 23% of full-length 16S rRNA sequences remained unclassified to the species level and that the resolution provided by the V1-V3 region is comparable with that of the full-length gene (13). Longer amplicons generally reduce PCR amplification efficacy, as has also been demonstrated for full *versus* partial amplification of the 16S rRNA gene (29). Additionally, primers for amplification of the full-length 16S rRNA gene may be less universal than well-established alternatives for partial amplification (20, 30, 31). In clinical microbiology, where samples often contain low levels of bacterial DNA with a background of abundant human DNA, PCR sensitivity and specificity should be prioritized over a questionable gain in species-level resolution. Therefore, here, a well-established real-time 16S rRNA gene V1-V3 PCR was used for ONT sequencing. The use of this segment simplifies the transition to ONT sequencing for clinical laboratories that have a validated real-time 16S rRNA gene V1-V3 PCR in place.

To increase species-level resolution, supplementing partial amplification of the 16S rRNA gene with partial *rpoB* amplification is helpful. With such an approach, the 16S rRNA gene amplification ensures maximal sensitivity, while the *rpoB* target provides improved species discrimination. In addition, as demonstrated here, the combination of two targets reduces the risk of missing species due to primer mismatches and increases detection rates for low-concentration species in complex polymicrobial scenarios. Replacing the 16S rRNA gene with *rpoB* is not advisable, as *rpoB* PCR is less efficient and shows more cross-reactivity with human DNA, leading to a lower sensitivity for weakly positive samples (18).

The RipSeq LRS module for ONT amplicon data is based on a two-step principle, with k-mer based mapping against a suitable database (binning), as a first step. Thereafter, a random selection of sequencing reads binned to a reference is used to generate a consensus sequence subsequently identified in a BLAST search against the same database, providing both homology with the best-matching species and distance to the next alternative. This approach mitigates problems related to the analysis of Nanopore amplicon data, as random sequencing errors are nullified through the consensus generation from each bin. Furthermore, when a minor proportion of reads from a given species are erroneously binned to a reference from another related species due to the Nanopore-associated errors and small 16S rRNA gene inter-species variations, the consensus sequence will still represent the biological sequence and be correctly re-assigned in the subsequent BLAST search. Finally, as demonstrated by the findings in Supplementary Table S4, the described approach allows for recognition of novel species or species not present in the database. For these, binning against the closest available reference in the database, followed by consensus generation, will result in a low score in the subsequent BLAST search. Such low-scoring consensus sequences can either be submitted to a BLAST search against a larger database (e.g., GenBank) or reported at a higher taxonomic level, depending on the homology obtained.

This study has several limitations. Except from the commercial mock community, we cannot know the true composition of the samples. Although 16S rRNA sequencing is a reference method in diagnostic bacteriology low abundance species in polymicrobial samples can, as discussed above, remain undetected for various reasons. Therefore, when comparing four amplicon-based approaches, we did not define a reference standard, and accepted all identifications not defined as background contamination as true in the clinical samples. The use of e.g., a composite reference standard requiring detection by at least two methods, would define relevant identifications of bacteria made by only one method as false positives, even when not suspected to represent background contaminant DNA. This would have masked the lower detection rate for low abundancy species in complex samples, leveled differences in detection rates and resulted in a lower calculated specificity for the most sensitive methods. Furthermore, multiplexing samples on the same MinION flow cell may be possible, but may introduce sample cross-contamination and bleeding, which were not assessed here. Preliminary data generated by the authors (not shown) suggest that this is not a substantial challenge.

In conclusion, the results presented demonstrate that Nanopore sequencing of partial 16S rRNA gene and *rpoB* amplicons directly from clinical samples is feasible, with results paralleling those of Illumina sequencing of the same genetic markers. Among the four target/platform combinations tested, Nanopore sequencing of the 16S rRNA V1-V3 provided the best sensitivity and improved species-level resolution compared to the V3-V4 segment used with Illumina. Including *rpoB* as a supplementary target enhanced both sensitivity and species-level discrimination for the two platforms. ONT offers shorter turn-around times enabling next-day results. Combined with a Windows/MacOS-based bioinformatics tool (RipSeq LRS), eliminating the need for a dedicated bioinformatician, the presented approach represents an attractive option for clinical laboratories.

## Data Availability

The 16S rRNA gene and rpoB sequences generated in this study will be deposited in the European Nucleotide Archive (ENA) at European Molecular Biology Laboratory-European Bioinformatics Institute (EMBL-EBI). We are in the process of submitting the data.

## ACKNOWLEDGEMENTS

This work was supported by the Western Norway Regional Health Authority’s research funding [grant number F-12826].

